# Cytokine release syndrome-like serum responses after COVID-19 vaccination are frequent but clinically inapparent in cancer patients under immune checkpoint therapy

**DOI:** 10.1101/2021.12.08.21267430

**Authors:** Thomas Walle, Sunanjay Bajaj, Joscha A. Kraske, Thomas Rösner, Christiane S. Cussigh, Katharina A. Kälber, Lisa Jasmin Müller, Sophia Boyoung Strobel, Jana Burghaus, Stefan Kallenberger, Christoph Stein-Thöringer, Maximilian Jenzer, Antonia Schubert, Steffen Kahle, Anja Williams, Birgit Hoyler, Lin Zielske, Renate Skatula, Stefanie Sawall, Mathias F. Leber, Russell Z. Kunes, Johannes Krisam, Carlo Fremd, Andreas Schneeweiss, Jürgen Krauss, Anne Katrin Berger, Georg M. Haag, Stefanie Zschäbitz, Niels Halama, Christoph Springfeld, Romy Kirsten, Jessica C. Hassel, Dirk Jäger, NCT ANTICIPATE Investigators, Guy Ungerechts

## Abstract

Cancer patients frequently receive immune checkpoint therapies (ICT) which may modulate immune responses to COVID-19 vaccines. Recently, cytokine release syndrome (CRS) was observed in a cancer patient who received the BTN162b2 vaccine under ICT. Here, we analyzed adverse events (AEs) in patients of various solid tumor types undergoing (n=64) or not undergoing (n=26) COVID-19 vaccination under ICT as an exploratory endpoint of a prospectively planned cohort study. We did not observe clinically relevant CRS after vaccination (95% CI [0,0.056]). Short term (<4 weeks) serious AEs were rare (12.5%) and overall AEs under ICT were comparable to unvaccinated patients. Despite the absence of CRS symptoms, we observed a pairwise-correlated set of CRS-associated cytokines upregulated in 42% of patients after vaccination and ICT (>1.5fold). Hence, clinically meaningful CRS appears to be rare in cancer patients under ICT and elevated serum cytokine levels are common but not sufficient to establish CRS diagnosis.

## Introduction

Patients with solid tumors have an increased fatality risk after infection with the SARS-CoV-2 coronavirus (Russell et al., 2021). Cancer patients have therefore been prioritized for vaccination against SARS-CoV-2 (COVID-19 vaccination) in many countries (Ribas et al., 2021; Trapani & Curigliano, 2021). Approved vaccines in Europe and the United States use mRNA lipid nanoparticles or viral vectors to transiently transfect/transduce a SARS-CoV-2 spike mRNA/transgene which is translated in the patient’s healthy cells at the site of vaccination, thus strongly inducing cellular and humoral adaptive immunity (Baden et al., 2020; Fendler et al., 2021; Folegatti et al., 2020; Frenck et al., 2021; Sahin et al., 2014). However, cancer patients were underrepresented in clinical phase III trials leading to FDA and EMA approval of these vaccines (Baden et al., 2020; Folegatti et al., 2020; Polack et al., 2020). Moreover, an increasing number of cancer patients receive immunomodulatory cancer therapies, mostly immune checkpoint therapies (ICT) blocking the PD-1/PD-L1 coinhibitory axis for T cell activation (Haslam & Prasad, 2019). Since ICT leads to reactivation of tumor antigen-reactive T cells, it is possible that ICT may also influence activation of SARS-CoV-2 spike protein (S1)-specific T cells. This increased T cell activation may lead to massive cytokine release and subsequent clinical reactions. The body’s systemic reaction to the resulting release of multiple inflammatory cytokines from T and myeloid cells is called cytokine release syndrome (CRS). CRS manifests itself in fever, hypotension, hypoxia and multiorgan dysfunction at later stages (Lee et al., 2019). Most frequently such responses are observed after adoptive T cell therapies, bispecific antibodies to the CD3 co-receptor or severe infection (Fajgenbaum & June, 2020). CRS is commonly graded according to the Common Terminology of Adverse Events (CTCAE) or the American Society for Transplantation and Cellular Therapy (ASTCT) consensus grading (Fajgenbaum & June, 2020; Lee et al., 2019). However, fever ≥38°C alone is sufficient to establish CTCAE grade 1 CRS, which does not account for mild fever as part of many appropriate immune reactions. Hence, an exhaustive differential diagnosis is essential in establishing CRS (Fajgenbaum & June, 2020).

Au et al. recently reported a patient with CRS without evidence for infection after COVID-19 vaccination under ICT. This patient required hospitalization due to fever, thrombocytopenia (grade 3 CTCAE 4.03, grade 1 ASTCT) as well as increased c-reactive protein levels (CRP > 200mg/l), prompting questions about the frequency of CRS in cancer patients under ICT. However, it remains unclear how frequently CRS occurs, and whether cytokine profiles could be explored to identify ICT-treated cancer patients at risk for CRS at early stages.

Here, we assessed adverse events, clinical laboratory data and serum cytokine responses in patients undergoing combined ICT and COVID-19 vaccination as an exploratory endpoint of a prospectively registered single center cohort study.

## Results

### A pan-tumor cohort study across diverse immune combination therapies

Between 2^nd^ December 2019 and 20^th^ July 2021, we screened 301 patients, of which 190 were screened prospectively and 111 retrospectively as defined in the trial protocol (German Clinical Trials Register, DRKS00022890). Of these patients, 29 patients with dermatological cancers were also included in a retrospective survey study at our center (Strobel et al, manuscript in revision). We recruited 220 patients with advanced solid tumors undergoing immune checkpoint therapy (ICT) at our center (Figure 1A). These patients received regular blood sampling before and during therapy and were monitored for adverse events. We assessed the COVID-19 vaccination status of the patients within the cohort during treatment follow-up and identified 64 patients who received a COVID-19 vaccine while under ICT and 26 patients who did not (Figure 1A, B). The remaining patients either died before the vaccine was widely available (n=52), did not have their vaccination status assessed (n=52), were lost to follow up (n=15), refused to disclose their vaccination status (n=5) or received COVID-19 vaccinations before ICT (n=6) (Figure 1A).

**Figure 1.**
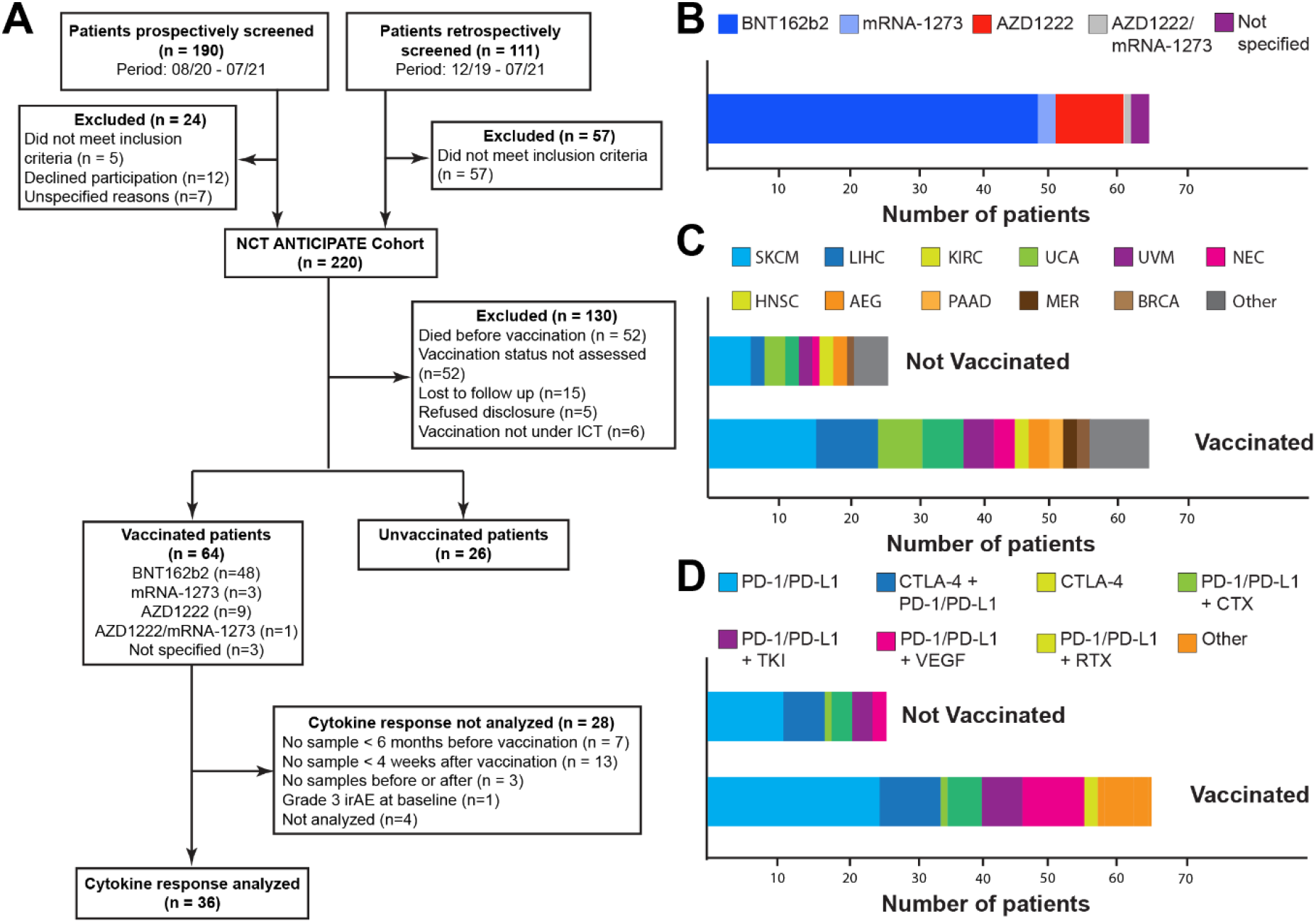
A pan-tumor cohort study across diverse immune combination therapies. (A) CONSORT flow chart indicating patient screening data and cohorts for subsequent data analysis. (B) Stacked bar-graph depicting the type of COVID-19 vaccination for vaccinated patients (n=64). (C) Stacked bar-graph indicating tumor types of vaccinated (n=64) and unvaccinated (n=26) patients. (D) Stacked bar-graph indicating immune combination therapies of vaccinated (n=64) and unvaccinated (n=26) patients.

We focused further analyses on the 90 patients with known vaccination status (Table 1). 23 cancer types were represented within this patient group, the most frequent being skin melanoma (n=21), hepatocellular carcinoma (n=11) and renal cell carcinoma (n=9) (Table 1, Figure 1C). Therapies included a variety of combinatorial immunomodulatory therapies, most frequently anti-PD-1/PD-L1 monotherapy (n=35), combined anti-PD-1 and anti-CTLA-4 therapy (n=15), and a combination of anti-PD-1/PD-L1 with anti-VEGF (n=11) (Table 1, Figure 1D). Despite the limited patient sizes of our cohorts, vaccinated and unvaccinated patients showed similar clinical characteristics such as sex, age, tumor type, stage, comorbidities, therapy regimen and line of therapy (Table 1) and were hence deemed suitable for further comparisons.

**Table 1.**
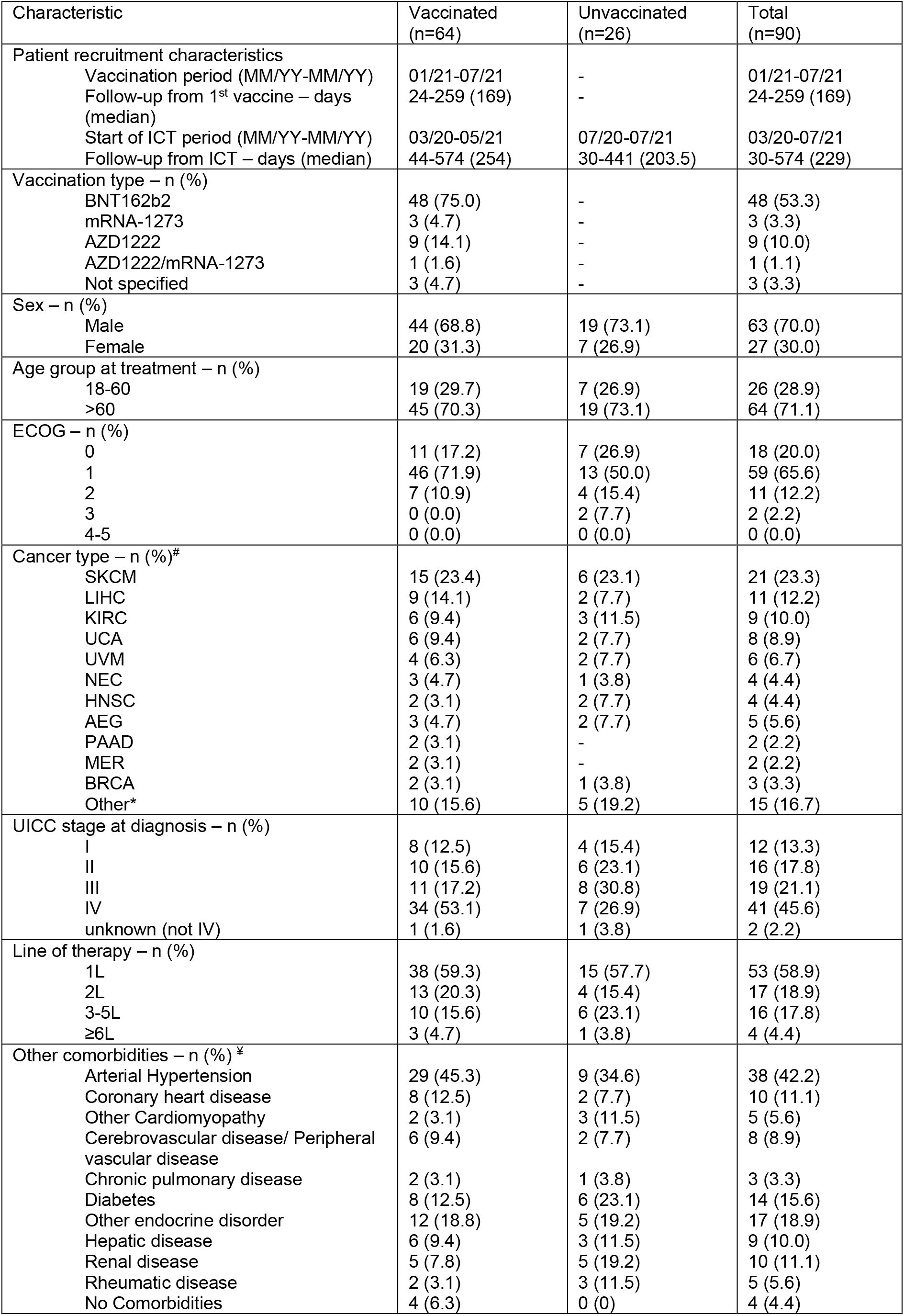
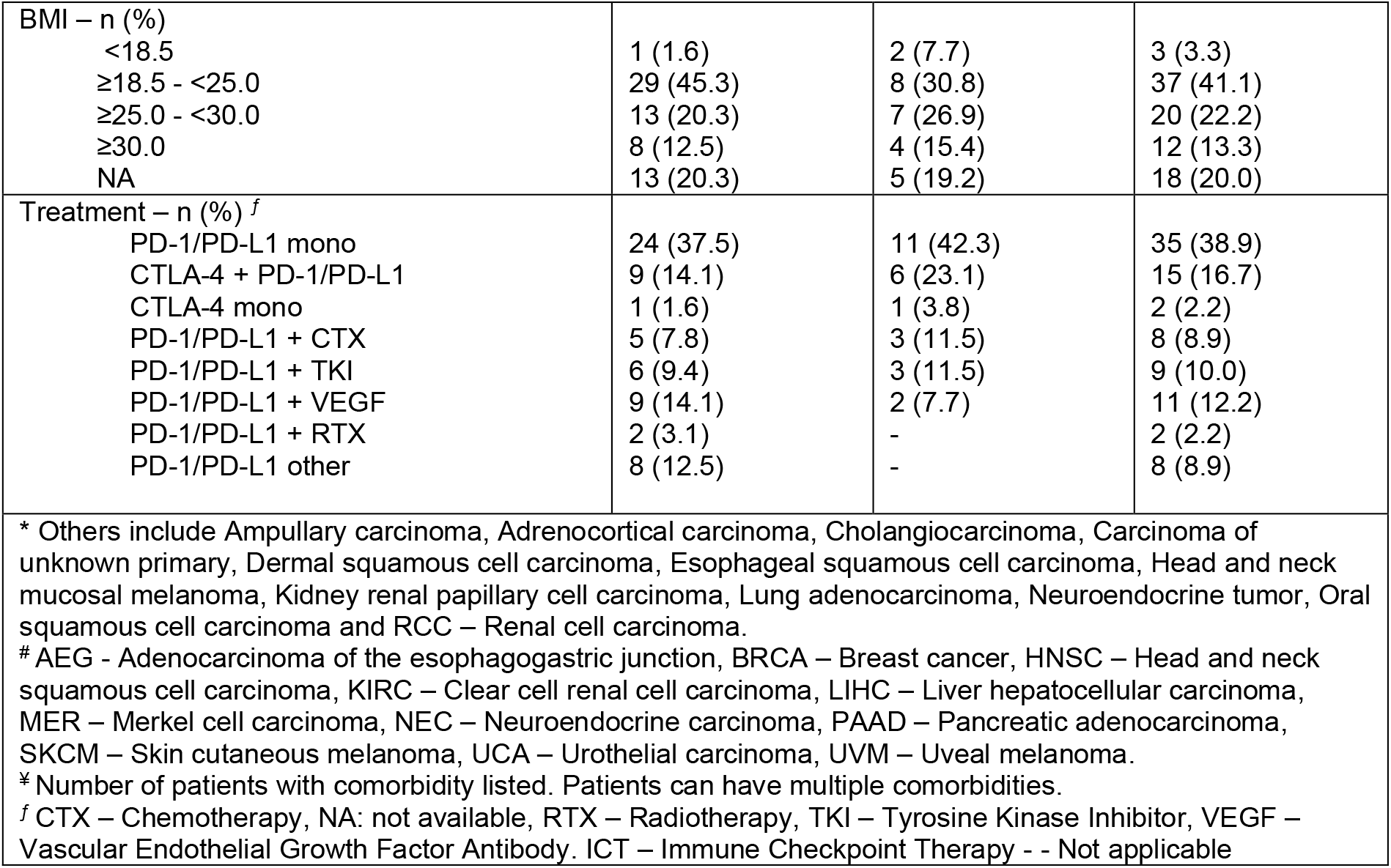
Characteristics of analyzed patients at baseline.

### Cytokine release syndrome is infrequent after COVID-19 vaccination under immune checkpoint therapy

To estimate safety of COVID-19 vaccination under ICT, we analyzed early adverse events (AE) from the first until 4 weeks after the second COVID-19 vaccination dose (Table 2, Figure 2A). While fewer early local adverse events such as pain at the injection site (n=2, 3.1%) were reported in our cohort, early systemic AEs were comparable to reported AEs in cancer patients including patients under ICT (Monin et al., 2021; Waissengrin et al., 2021). The most common systemic AEs included fatigue (n=10, 15.6%), muscle weakness (n=8, 12.5%) and fever (n=4, 6.3%) (Figure 2B, Table 2). Seven patients (10.9%) were hospitalized due to grade ≥3 AEs and three of these patients died (4.7%) (Figure 2B). One patient who received the BNT162b2 vaccine was admitted after grade 4 anemia due to esophageal varices bleeding and recovered quickly under high-dose proton pump inhibitor therapy. A second patient experienced grade 3 increase of transaminases under pembrolizumab + axitinib and the mRNA-1273 vaccine which normalized within three weeks after initial IV methylprednisolone and subsequent oral glucocorticoid tapering. Another patient already exhibited grade 2 diarrhea before BNT162b2 vaccination which worsened to grade 3 two weeks after vaccination. Multiplex-PCR analyses of stool showed Clostridium difficile, and symptoms improved after therapy with IV fluids and antibiotics. Another patient who received the AZD1222 vaccine was admitted due to grade 3 diarrhea for which no infectious cause could be determined and was therefore deemed ICT-related. Symptoms resolved under IV fluids and corticosteroids with oral tapering. Two patients were admitted after CT confirmed fulminant hepatic disease progression causing cholestasis and both patients ultimately died. Finally, one patient with a history of combined severe aortic stenosis (0,8cm^2^ aortic valve area) and aortic insufficiency died at home after cardiac decompensation with pleural effusion and lower limb edema after AZD1222 administration.

**Table 2.** Early adverse events after COVID-19 vaccination (<4 weeks)

| Adverse Event | Any grade | Grade ≥3 |
| --- | --- | --- |
| Any adverse event – n (%) | 40 (62.5) | 8 (12.5) |
| No adverse events – n (%) | 24 (37.5) | 62 (87.5) |
| Adverse event – n (%) |  |  |
| Fatigue | 10 (15.6) | 2 (3.1) |
| Pain at injection site | 2 (3.1) | 0 (0.0) |
| Muscle weakness | 8 (12.5) | 3 (4.7) |
| Arthralgia | 1 (1.6) | 0 (0.0) |
| Fever | 4 (6.3) | 0 (0.0) |
| Infection | 4 (6.3) | 3 (4.7) |
| Nausea | 2 (3.1) | 0 (0.0) |
| Headache | 2 (3.1) | 0 (0.0) |
| Vomiting | 1 (1.6) | 0 (0.0) |
| Upper GI bleeding | 1 (1.6) | 1 (1.6) |
| CRS grade ≥2 | 0 (0.0) | 0 (0.0) |
| Laboratory abnormalities - n (%) |  |  |
| Anemia | 7 (10.9) | 1 (1.6) |
| Thrombocytopenia | 8 (12.5) | 1 (1.6) |
| Bilirubin increase | 9 (14.1) | 1 (1.6) |
| Creatinine increase | 6 (9.4) | 1 (1.6) |
| Death – n (%) | 3 (4.7) |  |

**Figure 2.**
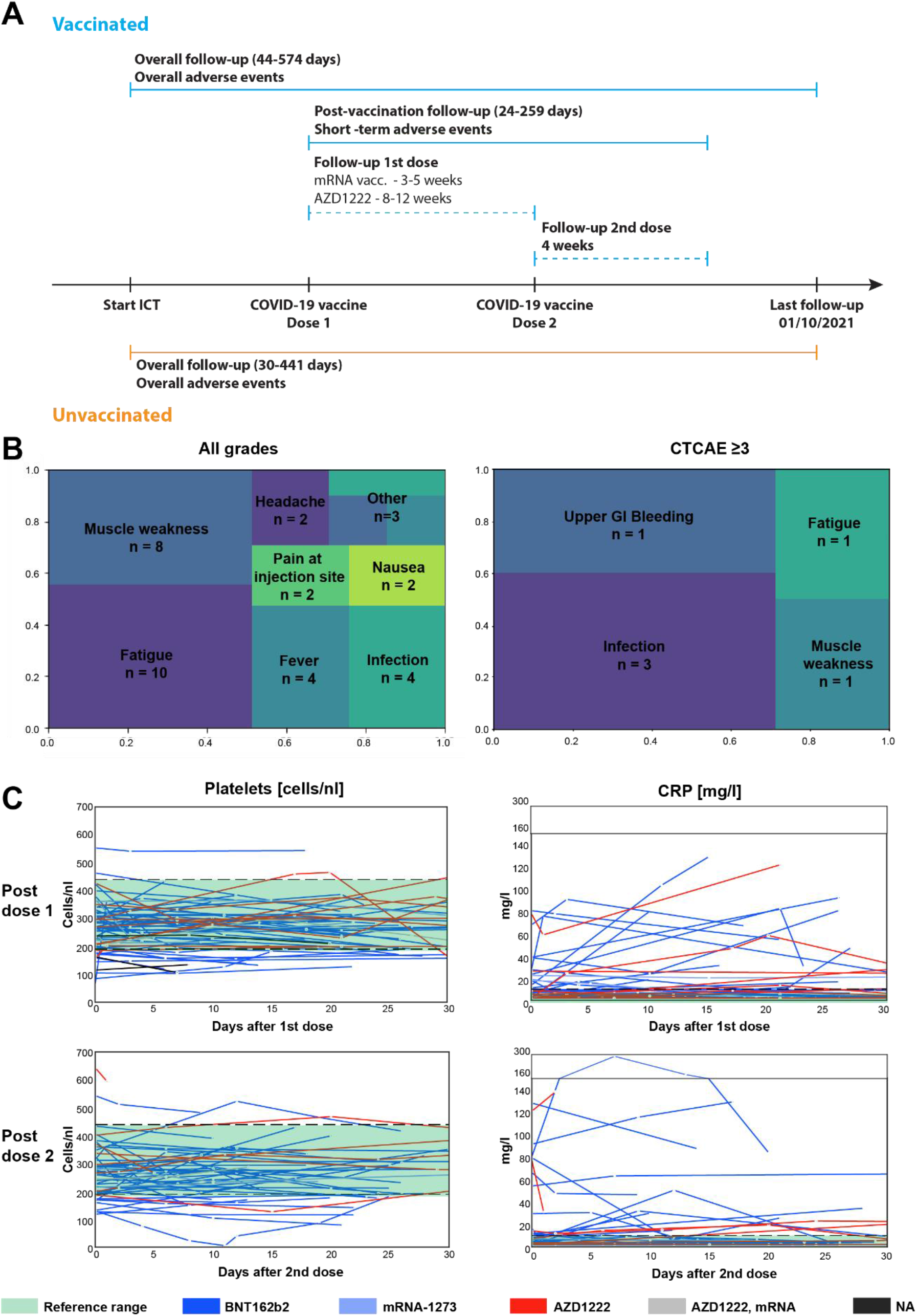
Early adverse events after COVID-19 vaccination under immune checkpoint therapy. (A) Schematic timeline depicting the start and duration of follow-up and vaccination timepoints of vaccinated and unvaccinated patients (B) Tree maps visualizing the proportion and the numbers of all and grade ≥3 AEs up to 4 weeks after vaccination in vaccinated patients (n=64). (C) Line-plots indicating platelet counts (n=61, left panel) and c-reactive protein concentrations ((CRP, n=55, right panel) either after the first vaccine dose (upper panels) or second vaccine dose (lower panels). Reference ranges are indicated in green shading. Lines are colored according to the vaccination schema used with the color code indicated below. NA: not available.

We observed no clinically relevant CRS in our patient cohort, defined as all CRS without evidence for infection identified by either the CTCAE v5.0 or ASTCT 2019 criteria (95% CI [0,0.056]). We excluded patients with fever alone without constitutional symptoms, which may reflect appropriate inflammatory reactions (Fajgenbaum & June, 2020). In the above-mentioned case report (Au et al., 2021) CRS was associated with thrombocytopenia and c-reactive protein (CRP) increase. In our cohort, only one patient experienced grade ≥3 thrombocytopenia with a platelet count of 5/nl four days after the 2^nd^ BNT162b2 dose. This patient had received gemcitabine and carboplatin three days prior to the event while still under prednisolone (50mg/d) due to a grade 3 autoimmune hemolytic anemia, which started after a blood transfusion two months earlier. The patient was asymptomatic, afebrile and was not hospitalized. Moreover, platelet counts spontaneously normalized within 2 weeks thus making CRS unlikely. We frequently observed mild (>30mg/l and >1.5-fold) CRP increase after vaccination (n= 22, 40% after 1^st^ dose; n=17, 35% after 2^nd^ dose) (Figure 2C). One patient showed a severe CRP increase (80 to 289mg/l) peaking 7 days after the 2^nd^ BNT162b2 dose (Figure 2C). This patient was also asymptomatic including absence of fever/hypotension or hypoxia, thus making CRS unlikely. Blood and urine cultures remained negative, and CRP spontaneously dropped below 100mg/l within 2 weeks. Hence, we did not observe any clinically apparent CRS after COVID-19 vaccination in our cohort suggesting that CRS may be rare in patients under ICT and concurrent COVID-19 vaccination.

### Asymptomatic CRS-like serum response patterns after COVID-19 vaccination under ICT

To evaluate cytokine responses indicative of CRS, we analyzed serum levels of CRS-associated cytokines in 37 patients undergoing concurrent ICT and COVID-19 vaccination with a baseline sample ≤6 months before vaccination and a sample ≤6 weeks after vaccination (Figure 3A-B). We excluded one patient who had an immune related adverse event (arthritis grade 3) at baseline before vaccination.

**Figure 3.**
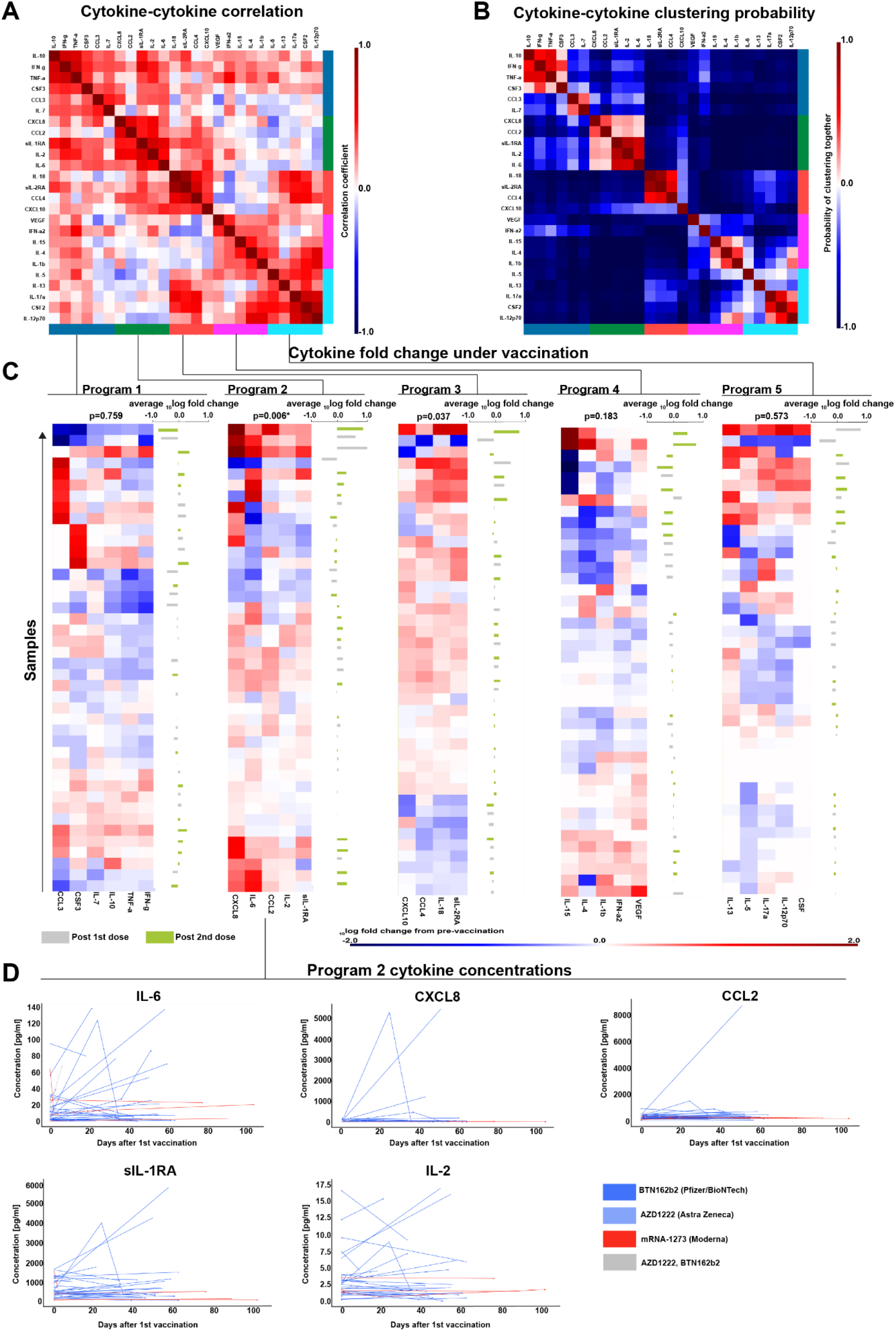
A correlated program of CRS-related cytokines is frequently upregulated after COVID-19 vaccination under immune checkpoint therapy. (A) Heatmap indicating spearman correlation indices of log(fold change) cytokine concentrations after COVID-19 vaccination from n=38 patients. Color bars on the sides indicate clusters obtained from hierarchical clustering. (B) Heatmap indicating probability of each pair of cytokines clustering together as calculated by bootstrapping (n=10000) from n=38 patients. (C) Heatmaps indicating _10_log-fold change of cytokine concentrations after vaccination. Bar graph on the side indicates average _10_log-fold change of cytokines in each row with concentrations after the first or second vaccination dose labeled according to the color code below from n=38 patients. P-values (two-tailed) were calculated using a Wilcoxon signed rank test. (D) Line-plots indicating cytokine concentrations of cytokine program 2 cytokines after vaccination from n=38 patients.

To analyze cytokines induced by vaccination under ICT, we performed pairwise correlation of all measured cytokines, which yielded four clusters of pairwise correlated cytokine programs (colored sidebars, Figure 3A). To assess the stability of this clustering we bootstrapped the probability of each pair of cytokines falling into the same cluster (see Methods). Bootstrapping confirmed the stability of cytokine programs 2 and 3 while the other clusters were more heterogeneous (Figure 3B). Assessment of the log fold change of cytokine concentrations indicated that cytokine programs 2 and 3 were upregulated after vaccination in most patients although only program 2 was statistically significant when correcting for multiple comparisons (p=0.006, q=0.03) (Figure 3C). Program 3 (p=0.04, q=0.09) included mediators previously described in CRS after the BNT162b2 vaccine in an anti-PD-1 treated cancer patient such as IL-18 and sIL-2RA (Au et al., 2021). Program 2 included hallmark CRS-cytokines and CRS-mediators indicative of T cell (IL-2) and myeloid cell activation (IL-6, CXCL8 = IL-8, CCL2, sIL-1R) (Figure 3D). IL-6 has been linked to CRS severity in multiple studies and surpassed 50pg/ml (research grade measurement) in 8 patients (22.2%), levels frequently observed in patients with severe COVID-19 or CAR-T cell induced CRS (Chen et al., 2021; Galván-Román et al., 2021; Hay et al., 2017) (Figure 3D). Stratification by patient characteristics revealed that cytokine program 2 was predominantly upregulated after BNT162b2 vaccination although only few patients received other vaccines in our study (Figure S1). Moreover, induction of cytokine program 2 was more frequent in male patients across a broad array of tumor types (Figure S1). Overall, our results suggest induction of CRS-related cytokines as a frequent but clinically inapparent event after COVID-19 vaccination in ICT treated cancer patients.

### Comparable adverse events in COVID-19 vaccinated and unvaccinated patients under ICT

To assess whether vaccination increased the frequency of ICT-related adverse events at later timepoints we compared AE frequencies in vaccinated (n=64) and unvaccinated patients (n=26) over the entire course of ICT. We did not detect any significant differences in all grade or grade ≥3 AEs between vaccinated and unvaccinated patients under ICT (Figure 4A-B). One patient experienced CRS grade 2 before COVID-19 vaccination but no patient showed CRS after vaccination. Immune related AE were numerically more frequent in unvaccinated patients while vaccinated patients had a higher frequency of fatigue, nausea and lower grade infections although these differences were not statistically significant (Figure 4A-B, Table 3). To confirm the accuracy of this comparison, we calculated propensity scores based on age and sex and matched vaccinated and unvaccinated patients according to this score (Figure S2A-B). For this purpose, we assigned unvaccinated patients a “virtual” vaccination date at the same time interval from the start of ICT as their vaccinated matched counterparts (Figure S2A). Again, overall and grade ≥3 adverse events were comparable between the matched cohorts (Figure S2C) suggesting that it is unlikely that COVID-19 vaccination profoundly increases the incidence of severe adverse events in ICT-treated cancer patients.

**Figure 4.**
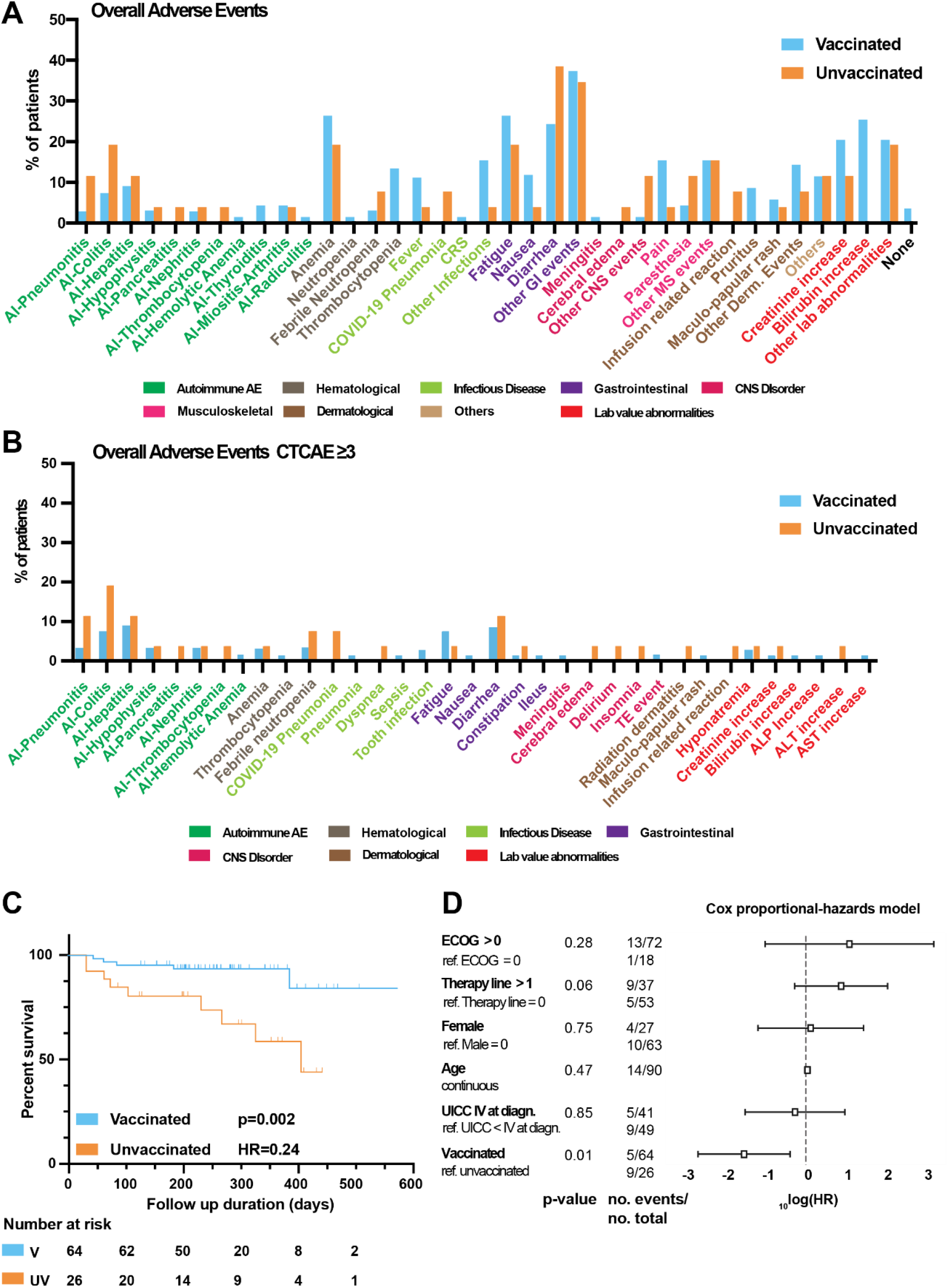
Comparable adverse events and prolonged overall survival in COVID-19 vaccinated immune checkpoint therapy treated cancer patients. (A-B) Grouped bar plots indicating the frequency of overall adverse event (A) or grade ≥3 adverse events (B) under immune checkpoint therapy in vaccinated (n=64) and unvaccinated (n=26) patients. (C) Kaplan-Meier curve indicating overall survival probability of vaccinated (n=64) and unvaccinated (n=26) patients under immune checkpoint therapy. P values and hazard ratios were calculated using a log-rank test. (D) Forest plot indicating the results of the Cox proportional hazards model of n=90 patients with boxes indicating the _10_log(hazard ratio) and whiskers indicating the 95% confidence interval. P values (two-tailed) and number (no.) of events are indicated on the left.

**Table 3.** Immune related adverse events under immunotherapy in vaccinated and non-vaccinated patients. Table indicating immune related adverse event frequencies with P values calculated using Fisher’s exact test.

| Characteristic – n (%) | Vaccinated (n=64) | Unvaccinated (n=26) | P value |
| --- | --- | --- | --- |
| Any AE | 62 (96.9) | 25 (96.2) | 0.99 |
| Grade $\geq 3$ | 31 (48.4) | 18 (69.2) | 0.10 |
| AI-Pneumonitis | 2 (3.1) | 3 (11.5) | 0.14 |
| AI-Colitis | 5 (7.8) | 5 (19.2) | 0.15 |
| AI-Hepatitis | 6 (9.4) | 3 (11.5) | 0.71 |
| AI-Hypophysitis | 3 (4.7) | 1 (3.9) | 0.99 |
| AI-Pancreatitis | 0 (0.0) | 1 (3.9) | 0.29 |
| AI-Nephritis | 2 (3.1) | 1 (3.9) | 0.99 |
| AI-Thrombocytopenia | 0 (0.0) | 1 (3.9) | 0.29 |
| AI-Hemolytic Anemia | 1 (1.6) | 0 (0.0) | 0.99 |
| AI-Thyroiditis | 3 (4.7) | 0 (0.0) | 0.55 |
| AI-Miositis/Arthritis | 3 (4.7) | 1 (3.9) | 0.99 |
| AI-Radiculitis | 1 (1.6) | 0 (0.0) | 0.99 |

Starting from 15^th^ October 2020, all patients were screened for COVID-19 at every therapy session generally every 1-4 weeks using a rapid antigen test fulfilling the quality criteria of the German Federal Institute for Vaccines and Biomedicines. We detected two COVID-19 cases in the unvaccinated cohort (7.7%, 95% CI [1.6%,22.5%]) which had to be hospitalized for severe pneumonia. One patient recovered and was able to resume therapy 6 weeks later but died two months after due to disease progression. The other patient died from COVID-19 pneumonia on the intensive care unit. We detected no COVID-19 cases in the vaccinated patient cohort, despite regular screening (95% CI [0,5.6%]). Accordingly, patient serum post vaccination neutralized SARS-CoV-2 S1 protein binding to recombinant human ACE2 in a competitive immunoassay (Figure S3). Hence, our results corroborate the increasing evidence that the here investigated COVID-19 vaccines have clinically meaningful efficacy in ICT treated cancer patients (Fendler et al., 2021; Monin et al., 2021).

To explore whether vaccination status was associated with oncological outcomes, we compared overall survival of vaccinated and unvaccinated patients (Figure 4C). Vaccinated patients showed prolonged survival as compared to unvaccinated patients (HR 0.24, p=0.002) (Figure 4C). This effect could not be explained by COVID-19 related deaths alone (Supplementary Table 1) but was stable in all relevant patient subgroups (Figure S4). Moreover, we confirmed this result in a Cox proportional hazards model where vaccination status was the strongest predictor of prolonged overall survival (coefficient -1.54, 95% CI [-2.69,-0.40], p=0.01) (Figure 4D). Hence, these data suggest an unexpected association of COVID-19 vaccination and prolonged overall survival in our patient cohort.

## Discussion

In a prospectively planned cohort study, we here describe a set of CRS-related cytokines commonly upregulated after COVID-19 vaccination in ICT treated cancer patients despite the absence of clinical CRS phenotypes. None of our patients showed evidence for clinically relevant CRS after vaccination suggesting that CRS is an infrequent event after COVID-19 vaccination under ICT. Moreover, comparison to unvaccinated patients suggested that COVID-19 vaccination does not profoundly increase the rate of immune related or grade ≥3 adverse events but may decrease the rate of COVID-19 infection in ICT treated patients.

A recent case report of CRS after vaccination in a colorectal cancer patient treated with anti-PD-1 ICT (Au et al., 2021) highlighted the insufficient evidence regarding vaccine-related adverse events in ICT treated cancer patients. Au and colleagues presented a patient with fever, thrombocytopenia, CRP increase and elevation of several cytokines including IFN-γ, sIL-2R, IL-18, IL-16, IL-10 after vaccination and ICT as compared to cytokine levels before initiation of ICT and vaccination (Au et al., 2021). In our study we did not observe any CRS (95%CI [0,5.6%]) after COVID-19 vaccination after both short-term (1^st^ dose until 4 weeks after 2^nd^ dose) and long-term follow-up (med. follow-up = 24 weeks after 1^st^ dose). Moreover, AE under immunotherapy were comparable in vaccinated and unvaccinated patients. Thus, CRS is likely an infrequent event under combined ICT and COVID-19 vaccination.

Despite the absence of clinically relevant CRS, we observed a set of CRS-related cytokines coordinately and commonly induced after COVID-19 vaccination under ICT. This included the CRS hallmark cytokine IL-6 and other CRS-related cytokines (CXCL8, IL-2, CCL2, sIL1-RA). Induction of IL-6 has been reported after mRNA-based lipoplex tumor vaccination and symptoms are generally mild and self-limiting (Sahin, Oehm, et al., 2020). Moreover, we found higher IL-2 levels after vaccination, which may be explained by T cell activation and preferable TH1 T cell polarization as shown in healthy adults vaccinated with BNT162b2 (Sahin, Muik, et al., 2020). Our patients also showed coordinated release of CCL2 and CXCL8 levels after COVID-19 vaccination which can be explained by the activation of myeloid cells by the mRNA loaded lipid nanoparticles of the BNT162b2 vaccine (Liang et al., 2017). While CCL2 and IL-2 were also reported to be induced in the above-mentioned case report of CRS in a mismatch repair deficient colorectal cancer (CRC) patient, the CRS hallmark cytokines IL-6 and CXCL8 levels remained largely constant in this patient (Au et al., 2021). Our study did not include a CRC or mismatch-repair deficient patient who received COVID-19 vaccination. It is possible that the clinical course observed by Au et al. is a CRC or mismatch repair deficiency specific effect given the distinct T cell inhibitory mechanisms in these tumors which may render T cells more responsive to PD-1 disinhibition (Au et al., 2021; Pelka et al.). Notably, one patient in our study experienced grade 2 CRS before any COVID-19 vaccination was administered, highlighting that CRS can occur independent of vaccination under ICT and may not necessarily be vaccine-related. This is particularly important in cancer patients in a palliative setting with limited treatment options, as CRS treatments such as glucocorticoids may impair ICT efficacy and deprive patients of an important treatment option (Iorgulescu et al., 2021; Maslov et al., 2021). Our results suggest that CRS-related cytokines are commonly induced after COVID-19 vaccination and not sufficient to establish the diagnosis of CRS. Clinically relevant CRS should therefore be diagnosed in symptomatic patients after an exhaustive differential diagnosis.

Two cases of severe COVID-19 (95% CI [1.6%,22.5%]) occurred in our unvaccinated cohort but none in our vaccinated patients. We observed induction of neutralizing antibodies after vaccination thus corroborating current evidence that COVID-19 vaccines may have meaningful activity in ICT-treated cancer patients (Fendler et al., 2021) (Strobel et al, manuscript in revision).

Vaccinated patients also showed increased overall survival (OS) in our study and vaccination status was an independent predictor of OS in a Cox proportional-hazards model. This observation cannot be explained by COVID-19 related mortality of unvaccinated patients alone. In unvaccinated patients only one in nine deaths was caused directly by COVID-19 and another one from tumor progression shortly after COVID-19 infection. Because patients underwent regular rapid antigen-testing (q1w-q4w) it seems unlikely that we missed a relevant number of COVID-19 cases. It is also possible that the small sample size of our heterogenous cohort has skewed the survival analysis despite the similarity of vaccinated and unvaccinated patients in many clinical covariates. Our results should therefore be validated in larger patient cohorts. Alternatively, patients with worse disease status and symptoms may be more hesitant and less likely to get vaccinated. This hypothesis is supported by the fact that unvaccinated patients experienced numerically more severe adverse events even though this difference was not statistically significant. This observation may also be a result of increased health awareness or higher compliance regarding oncological therapy in vaccinated patients, an outcome we did not assess in this study. Another possible explanation is that the cytokine boost induced by vaccination may have reinforced anti-tumor immunity. Specifically, IL-2 induction, as observed in our vaccinated patients, can break ICT resistance in subcutaneous murine models (Sun et al., 2019). Overall, the tested COVID-19 vaccines were linked to favorable outcomes in our study and may have meaningful clinical activity in ICT treated cancer patients.

Despite these important insights, our study also has several limitations which should be considered in its interpretation. Adverse events under SARS-CoV-2 vaccination were not the primary endpoint of this study. Therefore, sample size was not optimized for this endpoint and our trial is not powered to estimate the exact frequency of rare AE under ICT and COVID-19 vaccination. Moreover, AE were assessed upon presentation at our day clinic every 1-6 weeks and not at a standardized early timepoint as performed for randomized controlled vaccination trials (Polack et al., 2020). While serious AEs were generally reported instantly, lower grade AEs may be underreported due to recall bias. Finally, all serum cytokine and antibody titer analyses are research grade and absolute concentrations from this study should not be used to establish clinical diagnoses. Strengths of our analysis include the prospective design, prospective recruitment of most patients, long-term follow-up, broad array of cancer types and combination ICTs.

In summary, induction of CRS-related cytokines after COVID-19 vaccination is common in ICT-treated cancer patients, but generally clinically inapparent and hence not sufficient to define CRS. Our study supports current clinical practice of COVID-19 vaccination in cancer patients under ICT.

## Methods

### Clinical trial

Patients presented in this study are part of the exploration cohort of a prospectively registered cohort study (DRKS00022890). The exploration cohort consisted of 220 patients of which 166 were recruited prospectively and 54 retrospectively. Adult patients with advanced solid tumors starting a new cancer immunotherapy either as mono- or combination therapy except adoptive cell therapies were eligible for inclusion. Informed consent, hemoglobin levels ≥8g/dl when additional blood samples were obtained and measurable disease according to RECIST 1.1 were obligatory requirements for inclusion. Patients were followed up at least every 1-6 weeks depending on the treatment regimen. Adverse events (AE) were retrieved from electronic patient health records and graded according to CTCAE v5.0 as per trial protocol. Other metadata collected included age at time of informed consent, sex, tumor type, stage and histology, mutational status, sites of metastases, history of tobacco use, pre-existing health conditions, concurrent medication and survival. Pre-existing health conditions were obtained from electronic patient records and defined according to the Side Effect Resource (SIDER v4.1, http://sideeffects.embl.de). Patients also received regular (q1-q4w) clinical laboratory tests including creatinine, bilirubin, CRP, hemoglobin, platelet, and leukocyte counts. Additionally, patients received longitudinal blood samples for cytokine measurements before the start of therapy, within 1-7 weeks after therapy initiation and every 8-12 weeks under immunotherapy. COVID-19 vaccination status was assessed during regular follow-up. Patients did not undergo any additional screening for determination of vaccination status.

The primary outcome measure of the trial was prediction of radiological response which will be reported elsewhere. Secondary outcome measures included the serum proteome and peripheral blood immune cell composition overall, grade 3 adverse events as well as progression-free and overall survival. Patient health information is pseudonymized. The study size was defined by sample size estimation based on the primary outcome parameter.

The trial was conducted in accordance with the declaration of Helsinki in its current edition. The trial received institutional ethics review board approval at Ethics Commission I Medical Faculty Heidelberg, Heidelberg University (S-373/2020, S-207/2005,) and Ethics Commission II Medical Faculty Mannheim, Heidelberg University (2021-567). Trial personal was subject to medical confidentiality (§ 9 Abs. 1 MBO-Ä), the general data protection regulation (DSGVO) and the data protection act of the state of Baden-Württemberg (LSDG).

### Analysis of serum cytokines and neutralizing antibodies

Blood was collected either peripherally or via a port catheter in coagulation matrix containing serum tubes (#01.1602, Sarstedt, Germany). Samples were kept at room temperature until preparation (<24h). For serum preparation, tubes were centrifuged at 2500g for 10min at room temperature, and the upper phase was transferred into 500µl aliquots and stored at -80°C. For both cytokine and antibody analysis we only selected patients with baseline samples not older than 6 months prior to vaccination. For cytokine analysis we selected all samples obtained up to 4 weeks after the 2^nd^ vaccination dose. For antibody analysis we selected all samples until the end of follow-up.

Serum samples were thawed and immediately analyzed in duplicates using the Legendplex Cytokine Storm Panel 1 (741091, Biolegend, CA, USA), Cytokine Storm Panel 2 (741142, Biolegend, CA, USA) or SARS-CoV-2 Neutralizing Antibody Assay (741127, Biolegend, CA, USA) according to manufacturer’s instructions and analyzed on a BD FACS Canto II flow cytometer (BD, NJ, USA). Analyte concentrations were interpolated from a standard curve using 5 parameter logistic regression using Legendplex Software v 2021.07.01. Cytokine concentrations below the lower limit of detection were set to 0. The acquisition and processing of the raw data was performed by a clinician scientist who was blinded to the patients’ identity and metadata and who was not involved in downstream data analysis. Notably, the patients’ pseudonyms contained the temporal sequence of the samples.

### General data analysis

All data analysis was performed using Python 3 in a Jupyter notebook or Graph Pad Prism 9.2.0 (GraphPad Software Inc, CA, U.S.A.). All computer code is provided under (https://github.com/wallet-maker/ANTICIPATE_COVID-19.git). Plotting was done using the Matplotlib (3.4.3) and Seaborn (0.11.2) packages. Plots were arranged using Adobe Illustrator 2021 (25.2.2, Adobe Inc, San Jose, CA, USA).

### Bootstrapping clustering probabilities of cytokine-cytokine pairs

We transformed cytokine concentrations according to the following formula ct = 10log(c+1) with ct as the log-1p-transformed cytokine concentration and c as the raw cytokine concentration in pg/ml for all vaccinated patients. We then normalized all post vaccination log-1p transformed concentrations by subtracting the respective log1p transformed cytokine concentrations of the baseline sample. Based on these normalized concentrations, we then calculated a spearman cytokine-cytokine correlation matrix. We calculated pairwise Euclidian distances using scipy’s (1.7.2) scipy.spatial.distance.pdist function. Using scipy’s scipy.cluster.hierarchy.linkage function with the UPGMA method, we obtained the row and column linkages from the untransposed and transposed Euclidian distance matrix respectively and transformed these into flat clusters by applying scipy.cluster.hierarchy.fcluster function using a cophenetic distance of 1.6.

We sampled the normalized log1p transformed cytokine concentration dataframe object with replacement with the same sample size as the initial dataframe and repeated the above-mentioned procedure to obtain flat clusters. This sampling and clustering was repeated n=10,000 times. For each pair of cytokines we then counted their co-occurrence in a cluster and summed the values for all 10,000 separate clusterings dividing the counts for each pair of cytokines by 10 000 to obtain an approximation of the probability for each pair of cytokines to fall into the same cluster.

### Survival analysis

Survival was analyzed by Kaplan-Meier curves and Log-rank tests (Mantel-Cox) using GraphPad Prism and the lifelines package (0.26.3). We inspected the Kaplan-Meier curves and did not see any obvious violation of the proportional hazards assumption. Using the lifelines package, we also calculated a Cox-proportional hazards model using sex, age at trial inclusion, stage, ECOG and vaccination status. We excluded BMI, tumor type and therapy type. BMI was not available for several patients which would have decreased the population size. When including therapy type and tumor type the Cox model did not converge, likely due to the large number of categories for these variables. When no events were observed in one group we reported hazard-ratios using the Mantel-Hanszel method implemented in Graph Pad Prism 9.2.0. Otherwise, we used the logrank method implemented in Graph Pad Prism 9.2.0.

### Propensity score matching

Propensity score matching was performed using the pymatch package (0.3.4). Propensity scores were calculated for each patient based on age and sex. Based on these propensity scores we assigned each unvaccinated patient a vaccinated counterpart with replacement. This led to efficient matching, defined as a reduction in the age and sex imbalance of the vaccinated and unvaccinated cohorts (Figure S2B). We did not include other variables into the calculation of the propensity score because they led to inefficient matching with greater imbalance of the cohorts after matching.

After matching the patients, we calculated the interval from start of immunotherapy to the first COVID-19 vaccination for each vaccinated patient. By adding this interval to the start date of immunotherapy of the respective matched unvaccinated patient we created a ‘virtual’ vaccination date for each unvaccinated patient. We then compared adverse events of vaccinated and unvaccinated patients after this (virtual) vaccination date.

### Statistical analysis and estimation of CRS frequency

Confidence intervals for CRS frequencies were calculated as Clopper-Pearson intervals based on the beta distribution using the statsmodels.stats.proportion.proportion_confint function of the statsmodels package (0.10.2). P values were calculated using Wilcoxon one sample tests or Wilcoxon matched-pairs signed rank tests for continuous/ordinal one sample or paired two sample data respectively using scipy’s scipy.stats.wilcoxon function (1.7.2). For categorical data/contingency tables we used Fisher’s exact test (GraphPad Prism 9.2.0). All p values are two-tailed. For cytokine data analysis p values were corrected for multiple comparisons with the Benjamini&Hochberg method using R 4.1.1 and the p.adjust function. For clinical data analysis we did not use multiple comparisons correction to increase our power to detect differences vaccine-related adverse events (which we did not find).

## Supporting information

STROBE_checklist

## Data Availability

All relevant data and code is supplied with the manuscript (https://github.com/wallet-maker/ANTICIPATE_COVID-19.git). Patient material of patients who have given consent for further analyses can be requested after approval of Ethics Commission I, Medical Faculty Heidelberg, Heidelberg University. No custom materials have been generated during this study.

## Author contributions

Conceptualization: T.W. Methodology: T.W. J.A.K., R.K.

Computer Code: T.W.

Investigation: T.W., S.B., J.A.K., B.H., L.Z., R.S., M.W., S.P.,

Patient recruitment: T.W., K.K., C.C., S.S, T.R, L.J.M., J.B., M.J., C.F., S.K., A.S., K.S., S.K., C.S-T., C.S., J.B., T.D., E.B., M.C., X.Z., A.R., M.S., F.M., M.L., A.S., D.J., J.K., G.M.H., S.Z., A.B., N.H., R.K., J.H.

Writing - Original Draft: T.W.,

Writing – Review and Editing: T.W., S.B., G.U., J.A.K., G.M.H, A.B.,C.S.

Visualization: T.W., S.B.

Funding Acquisition: T.W., G.U.

Supervision: G.U.

Statistical consultation: J.K., R.K.

## Acknowledgements

We thank all participating patients and their relatives for their consent to this research. We thank the NCT Day Clinic 1, Day Clinic 2 and Dermatology Outpatient Clinic nursing staff for blood sampling. We thank DKFZ Flow Cytometry Core Facility for technical support and infrastructure. T.W. and A.S. have been funded by a fellowship of the DKFZ Clinician Scientist Program, supported by the Dieter Morszeck Foundation.

---

**Figure S1.**
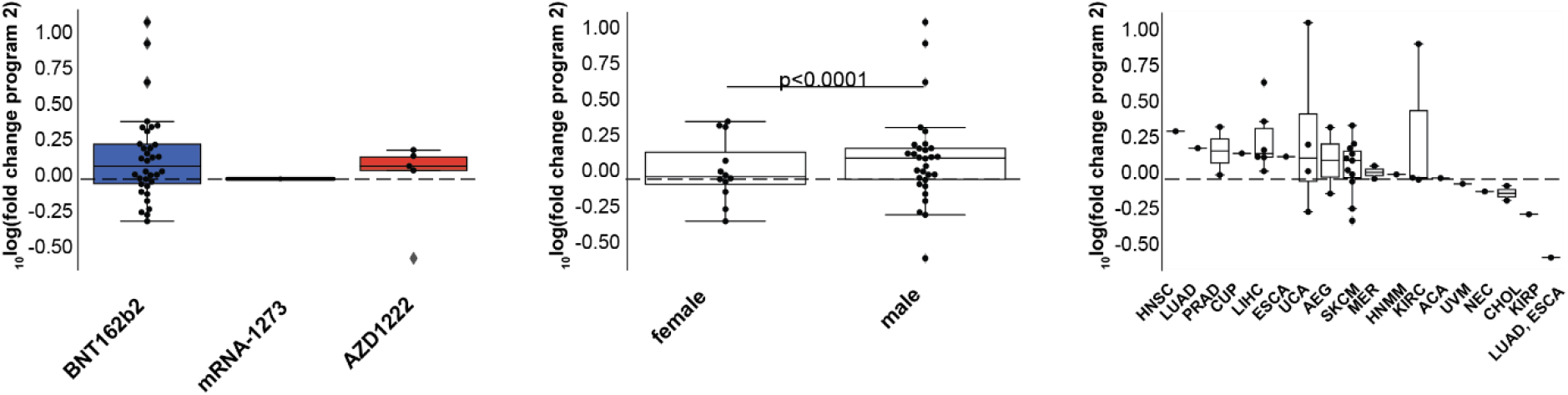
Cytokine program 2 upregulation stratified by patient characteristics. Box-plots indicating average _10_log(fold change) of cytokine program 2 after COVID-19 vaccination stratified according to vaccine type (left panel), sex (middle panel) or tumor type (right panel, n=38). P-value was calculated using a Mann-Whitney U-test.

**Supplementary Table 1.** Cause of death in vaccinated and non-vaccinated patients. Table indicating causes of death in vaccinated and unvaccinated patients. PD - progressive disease.

| Patient vaccination status | Cause of death |
| --- | --- |
| Unvaccinated | COVID-19 Pneumonia |
|  | Pulmonary metastases PD, respiratory insufficiency |
|  | Pulmonary metastases PD, respiratory insufficiency |
|  | Laryngeal PD, respiratory insufficiency |
|  | Respiratory Insufficiency |
|  | Intracerebral metastasis hemorrhage |
|  | Liver metastases PD, liver failure |
|  | Kidney failure |
|  | Unknown, patient declined therapy continuation |
| Vaccinated | Cardiopulmonary decompensation |
|  | Aortic stenosis, cardiopulmonary decompensation |
|  | Hepatic metastases PD, liver failure |
|  | Hepatic metastases PD, liver failure |
|  | Cholangiosepsis |

**Figure S2.**
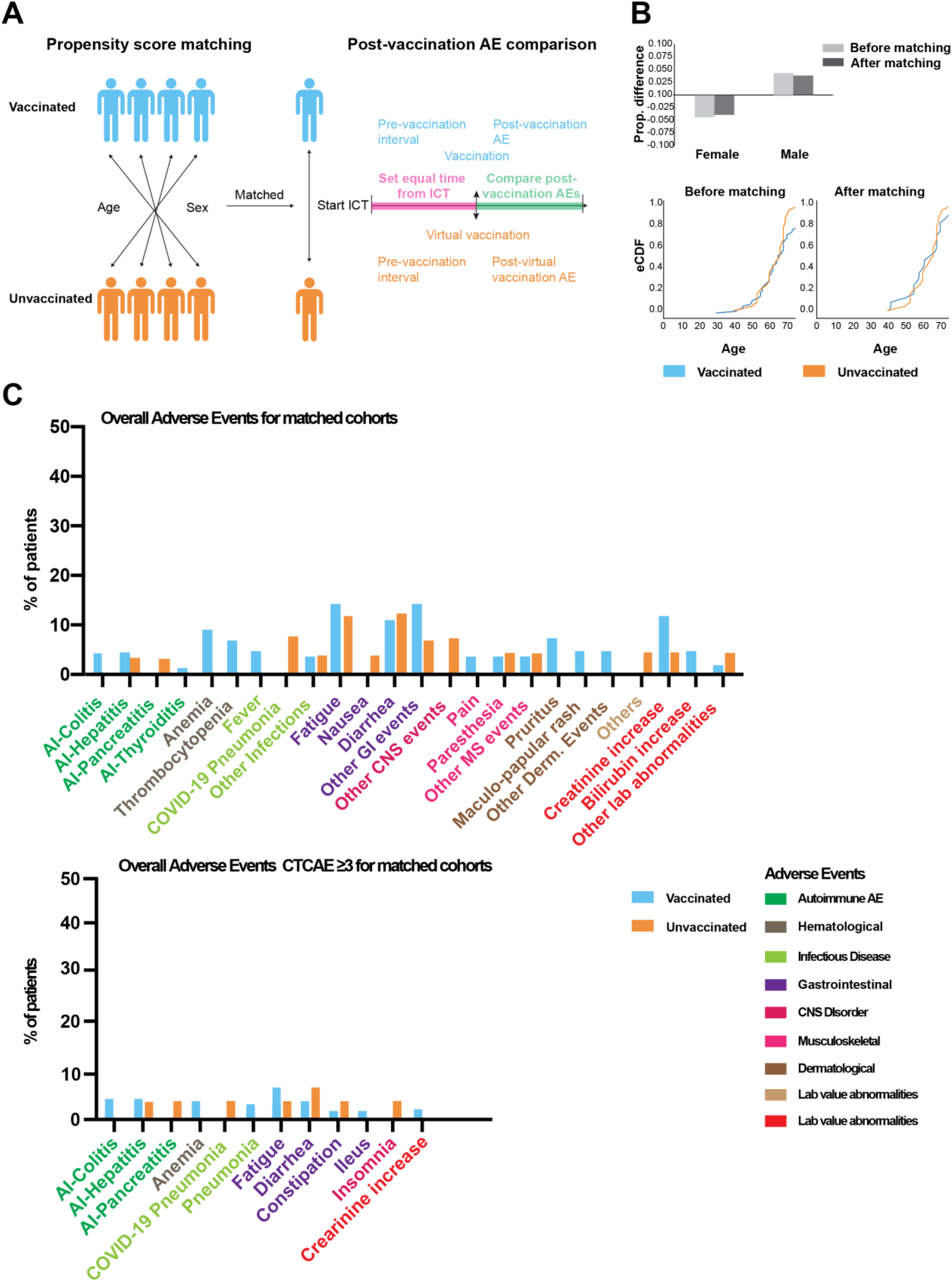
Comparable adverse events in sex and age matched COVID-19 vaccinated and unvaccinated immune checkpoint therapy treated cancer patients. (A) Schema indicating the propensity score based matching procedure (B) Box plots indicating the proportional difference of sex before and after propensity score matching (upper panel). Xy plot indicating the empirical cumulative distribution function of vaccinated and unvaccinated cohorts before and after matching. (C) Bar graphs indicating frequencies of all (upper panel) or grade ≥3 (lower panel) adverse events post (virtual) vaccination in matched vaccinated and unvaccinated patients.

**Figure S3.**
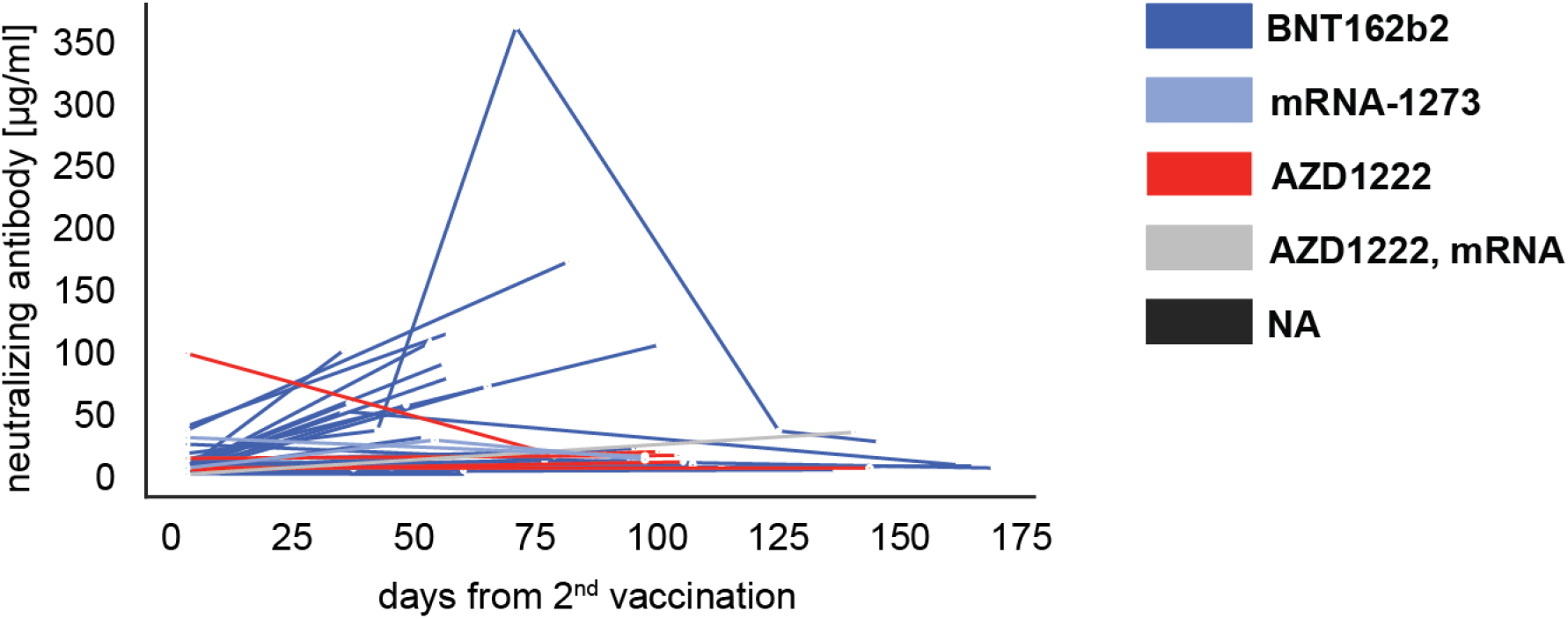
S1 neutralizing antibody concentrations after COVID-19 vaccination in ICT-treated patients. Line-plot indicating neutralizing antibody concentrations of individual patients after vaccination as assessed by an ACE2 S1 competitive immunoassay (research grade). Color code on the right indicating vaccine used in the respective patients.

**Figure S4.**
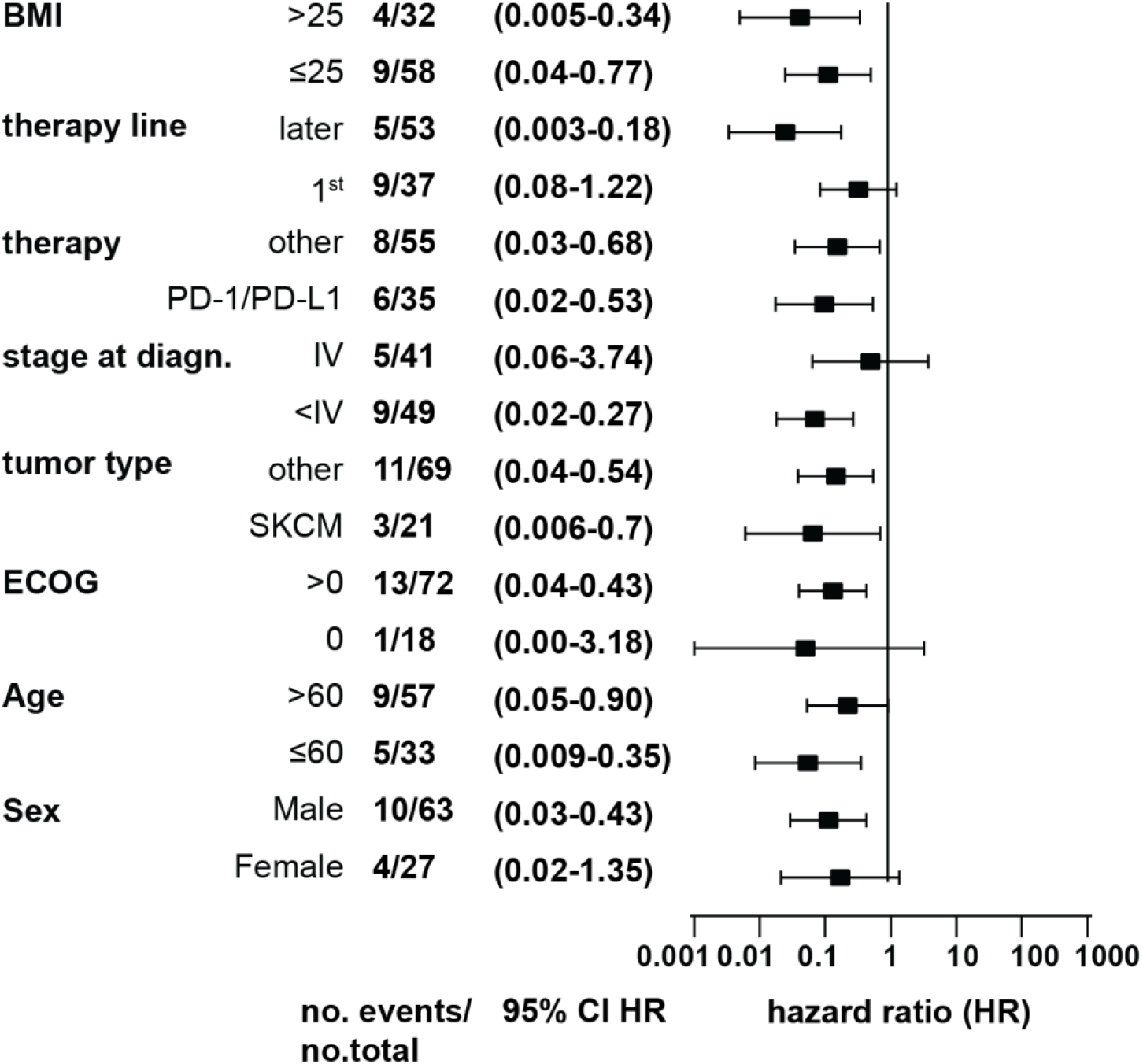
Vaccination is associated with of overall survival probability across patient subgroups. Forest-plot indicating hazard ratios (HR, boxes) and 95% confidence intervals (CI, whiskers) of vaccinated and unvaccinated patients stratified into the subgroups indicated with the exact number (no.) of events and patients at risk for each subgroup as well as the 95% CIs indicated on the left. Hazard ratios and confidence intervals were calculated using a Log-rank test or Mantel-Hanszel method if no events were observed in one group. Diagn.: diagnosis.

